# Feasibility, Acceptability, and Effectiveness of Group Antenatal Care on the Continuum of Care and Perinatal Outcomes in Sub-Saharan Africa: A Systematic Review and Meta-analysis Protocol

**DOI:** 10.1101/2024.09.22.24314166

**Authors:** Meresa Berwo Mengesha, Tesfaye Temesgen Chekole, Hiluf Ebuy Abraha, Etsay Weldekidan Tsegay, Abadi Hailay Atsbaha, Mihretab Gebreslassie, Zenawi Hagos Gufue

## Abstract

**Introduction:** In sub-Saharan Africa, the predominant model of individualized, one-on-one antenatal care has not significantly improved perinatal outcomes. Although the benefits of group antenatal care have been demonstrated in developed countries, its feasibility, acceptability, and effectiveness in resource limited settings, particularly in sub-Saharan African countries, have yet to be fully investigated. However, pilot studies show promising evidence of its effectiveness in these areas. This systematic review and meta-analysis will, therefore, review and summarize available studies and provide comprehensive and robust evidence that tends to contribute to the ongoing efforts to implement group prenatal care models in low-resource settings.

**Methods and analysis:** This systematic review protocol adheres to the Preferred Reporting Items for Systematic Reviews and Meta-Analysis Protocols guidelines. A comprehensive literature search will be conducted across multiple electronic databases, including PubMed/MEDLINE, Web of Science, EMBASE, and CINHAL, to identify pertinent articles published from January 1, 2016, to June 30, 2024. Experimental studies (pre-post, quasi-experimental study, cluster randomized controlled trial), prospective cohort design, prospective comparative study, and qualitative and mixed method designs will be included in the review. Abstract and full-text screening will be conducted by three reviewers using Covidence, according to the eligibility criteria set. The Joanna Briggs Institute (JBI) Critical Appraisal Tools, specifically designed for JBI Systematic Reviews, will be utilized to assess the methodological quality of the included studies. Statistical heterogeneity will be assessed using the Higgins test. Meta-analysis will be performed using R version 3.6.1 software and STATA version 16; applying random effects models to determine the weights. Pre-specified subgroup analysis and sensitivity analysis will be conducted as necessary. The study results will be reported sequentially, beginning with the primary outcomes, followed by secondary outcomes, and important subgroup analyses.

**Ethics and dissemination:** Ethical approval is not applicable as no original data will be collected. The findings of this review will be disseminated through publication and conference presentations.

**PROSPERO registration number CRD42024565501.**

## Introduction

Antenatal care (ANC) uptake is a crucial indicator for evaluating progress towards improving maternal outcomes (1). The provision of high-quality, women-centered ANC is especially crucial in low- and middle-income countries (LMICs), where pregnancy and perinatal outcomes are often disproportionately poor (2, 3, 4). In LMICs, particularly in sub-Saharan Africa, the predominant model of individualized, one-on-one care has not significantly improved perinatal outcomes (5). In contrast, group ANC (G-ANC) has emerged as a viable alternative service delivery model in high-income countries, linked to increased attendance, improved satisfaction, and better health outcomes for pregnant women and newborns (3, 6, 7). This model is also shown to benefit marginalized women, whose perinatal outcomes are comparable to those in developed countries (8, 9).

The World Health Organization (WHO) recommends G-ANC as an alternative to individual ANC, based on rigorous research and its contextual guidance promoting community mobilization through facilitated participatory learning and action cycles (9). The G-ANC is a transformative service delivery model that provides care to groups of eight to twelve pregnant women who are at similar gestational ages through cascades of scheduled meetings (10). This model incorporates physical assessments, education, skill development, and peer support and takes a more holistic, woman-centered approach in contrast to traditional ANC (11).

The Global G-ANC Collaborative acknowledges that it is crucial to adapt G-ANC models to the unique local contexts and priorities of LMICs to guarantee ownership, sustainability, and expansion (10). However, several challenges hinder the implementation of G-ANC in various settings, including recruiting and retaining participants, inadequate training and resources, a lack of focus on individual needs, financial barriers, and poor access to healthcare (12, 13, 14). These challenges can be overcome by developing effective recruitment and retention strategies, utilizing mixed methods to assess fidelity and investigate the potential of G-ANC-facilitated community groups, and implementing cost-effective measures (15, 16, 17).

Various studies have demonstrated that, as compared to the individualized care model, G-ANC is associated with increased attendance at ANC visits, improved quality of care, higher rates of facility-based deliveries, enhanced health literacy and client satisfaction, increased uptake of family planning methods, better birth weights, and higher rates of breastfeeding initiation and duration (18, 19, 20). Although the benefits of G-ANC have been demonstrated in developed countries, its feasibility, acceptability, and effectiveness in LMICs, particularly in sub-Saharan African, have yet to be fully researched. However, individual studies have reported encouraging results regarding the model’s efficacy in these areas, indicating the potential of G-ANC to enhance maternal and neonatal health outcomes in low-income settings (21, 22).

Therefore, a comprehensive and robust review of these studies is of paramount importance to evaluate the ongoing efforts to implement G-ANC models in diverse socio-economic contexts. This review defines G-ANC as the combination of conventional antenatal assessments with group discussions and support. The objective of this review is to synthesize the available evidence on the feasibility of G-ANC service delivery models in low-resource settings, its effectiveness of G-ANC in increasing ANC retention and facility-based deliveries, and its acceptability among pregnant women and community health workers. Additionally, this review aims to determine the overall impact of G-ANC on perinatal outcomes and the utilization of other maternal health services in sub-Saharan African countries.

## Methods

### Eligibility Criteria

#### Population

This review focuses on adult and adolescent pregnant women of diverse vulnerable populations in low-resource settings who are disadvantaged, marginalized, have normal or adverse pregnancy outcomes, have limited access to quality health care, and live in sub-Saharan Africa.

#### Exposure

G-ANC and/or Participatory Group

#### Comparator

For this review, the reference group comprises women whose ANC was provided by individualized, standard, conventional, or traditional models of care.

#### Outcomes

##### Primary outcomes

The effectiveness of G-ANC in increasing ANC retention and facility-based deliveries, its effect on perinatal outcomes (such as birth weight, preterm birth, neonatal intensive care unit admission, gestational age, maternal morbidity, and death), its influence on the utilization of other maternal health services (including family planning, postnatal care, assisted delivery, breastfeeding initiation, and duration).

##### Secondary outcomes

The feasibility of G-ANC service and its acceptability by pregnant women and community health workers will be considered secondary outcomes.

##### Setting and language

This review will focus on settings in sub-Saharan Africa, considering publication from January 1, 2016, to June 30, 2024, and publications published only in English.

##### Study design

Experimental studies (pre-post, quasi-experimental study, cluster randomized controlled trial), prospective cohort design, prospective comparative study, and qualitative and mixed method approach studies will be included in the review.

##### Exclusion

This review will exclude preprints, unpublished reviews, case reports, case series, commentaries, editor’s letters, and non-English publications.

##### Information sources

A comprehensive literature search will be conducted in various electronic databases, including PubMed/MEDLINE, Web of Science, EMBASE, and CINHAL, to identify pertinent articles. Information sources will be updated before submission to ensure all relevant studies are included. To guarantee the inclusion of all relevant studies, the reference lists of selected studies and systematic reviews with a similar scope will also be scanned.

##### Search strategy

The search strategy for identifying relevant literature was constructed using a combination of Medical Subject Headings (MeSH) and keyword terms that pertained to both the exposure and outcome of interest. Specific criteria were applied to the literature search, including restrictions on the date, language, and location of publication. Table 1 displays the pilot search strategies for PubMed and MEDLINE. The search will be further refined to target articles published in sub-Saharan Africa.

**Table 1.** Pilot search in the PubMed/MEDLINE database, June 30, 2024

| Database | Search restriction | Search strategy | # |
| --- | --- | --- | --- |
| PubMed/<br>MEDLINE | From January 1, 2016, to June 30, 2024<br>English<br>Exclude preprints | **#1 AND**<br>("Group antenatal care"[Text Word]) OR ("who antenatal care"[Text Word]) OR ("antenatal care services"[Text Word]) OR ("prenatal care"[Text Word]) OR ("participatory group"[Text Word]) OR ("positive pregnancy experiences"[Text Word]) NOT ("traditional antenatal care"[Text Word]) NOT ("individual antenatal care"[Text Word])<br>**#2 AND**<br>("Impact"[Text Word]) OR ("effect"[Text Word]) OR ("effectiveness of care"[Text Word]) AND ("continuum of care"[Text Word]) OR ("continuity of care"[Text Word]) OR ("utilization of antenatal care"[Text Word]) OR ("antenatal care attendance"[Text Word]) OR ("facility delivery"[Text Word]) OR ("quality of care"[Text Word]) OR ("postnatal attendance"[Text Word]) AND ("maternal self-efficacy"[Text Word]) OR ("maternal outcomes"[Text Word]) OR ("fetal outcomes"[Text Word]) OR ("perinatal outcomes"[Text Word])<br>**#3 AND**<br>Angola* OR Benin* OR Botswana* OR "Burkina | 111 |
|  |  | Faso*" OR Burundi* OR "Cape Verde*" OR<br>Cameroon* OR "Central African Republic*" OR<br>Chad* OR Comoros* OR Congo* OR<br>"Democratic Republic of Congo*" OR "Republic<br>of Cote d'Ivoire*" OR "Equatorial Guinea*" OR<br>Eritrea* OR Eswatini* OR Swaziland* OR<br>Ethiopia* OR Gabon* OR Gambia* OR Ghana*<br>OR Guinea* OR Guinea-Bissau* OR Kenya* OR<br>Lesotho* OR Liberia* OR Madagascar* OR<br>Malawi* OR Mali* OR Mauritania* OR<br>Mauritius* OR Mozambique* OR Namibia* OR<br>Niger* OR Nigeria* OR Rwanda* OR "Sao Tome<br>and Principe*" OR Senegal* OR Seychelles* OR<br>"Sierra Leone*" OR Somalia* OR "South Africa*"<br>OR "South Sudan*" OR Sudan* OR Tanzania*<br>OR Togo* OR Uganda* OR Zambia* OR<br>Zimbabwe* OR "West Africa*" OR "southern<br>Africa*" OR "south Africa*" OR "east Africa*"<br>OR Africa* OR "sub-Saharan Africa*" |  |

#### Protocol

The protocol for this systematic review was reported following the Preferred Reporting Items for Systematic Reviews and Meta-Analysis Protocols (PRISMA-P) guidelines (23). This review was registered on PROSPERO with registration number of CRD42024565501.

#### Patient and public involvement

Patients and/or the public were not involved in the design, conduct, report or dissemination plan of this research

#### Data management

The findings retrieved from the literature search will be imported, screened, and analyzed using professional software platforms such as STATA, R Studio, and Covidence. Covidence will facilitate the automatic elimination of duplications, coupled with a manual verification process for identifying resemblances among studies (e.g., publication year, authorship, journal details, etc.) conducted by the authors.

#### Selection process

Three reviewers (MG, ZHG, and HEA) will conduct abstract and full-text screening according to the eligibility criteria using Covidence. Any discrepancies among the three reviewers will be resolved with the assistance of other reviewers (MBM). A separate section will be set up in Excel specifically for relevant studies identified during full-text screening, which will include examination of reference lists of included studies and systematic reviews within the same domain. Rationales for exclusion will be meticulously recorded throughout the full-text screening process. A conclusive determination concerning the inclusion of studies will be reached by considering the outcomes of both Covidence and the spreadsheet. The presentation of study selection results will be facilitated by employing PRISMA flow diagrams **(Figure 1)**.

**Figure 1:**
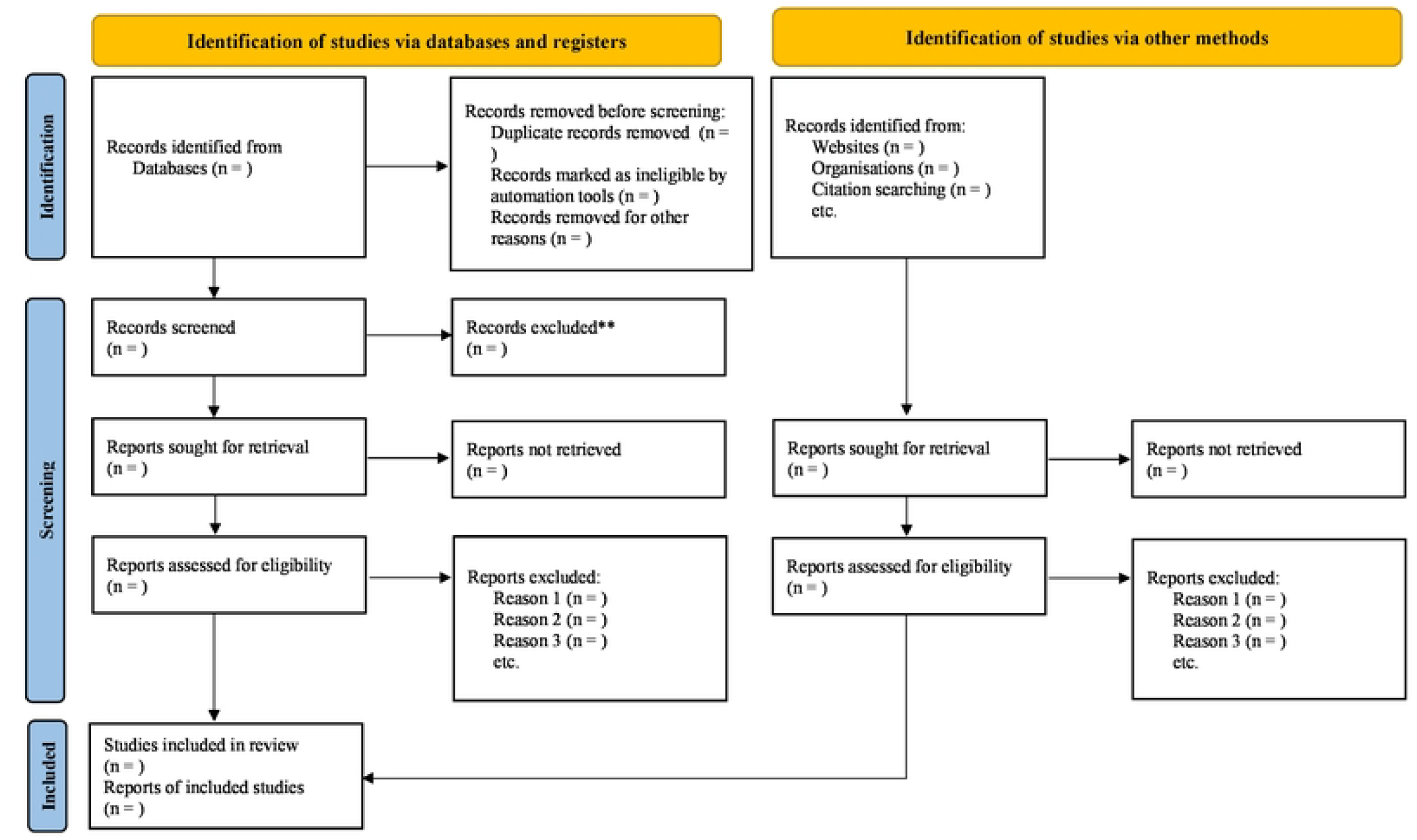
PRISMA 2020 flow diagram for new systematic reviews which included searches of databases, registers and other sources

#### Data collection

A standardized data collection tool will be developed in the form of a data extraction form. Subsequently, this form will undergo a pilot test among various groups, with potential modifications being made based on feedback received. Three reviewers (EWT, AHA, and TTC) will independently extract data from the studies included in the analysis. To ensure consistency in assessment methods, the reviewers will undergo a calibration exercise. In cases of discrepancies between reviewers, the principal investigator (MBM) will be consulted for resolution.

#### Data items

The following data items will be extracted from included studies: (1) Study data: title, author name, year of publication, country of study, journal, sample size, study period, study design, follow-up period (for experimental and cohort) and limitations; (2) Population: participant characteristics (number of group of women, number of women per group, how women are grouped, number of leaders in the group, who leads the group, the total number of group sessions (ANC sessions), length of the group session, etc.); (3) Intervention: G-ANC, or participatory group, involves assessing their experience, perspective, and outcomes within the group settings. (4)Comparison: the comparator used in the studies is conventional care or individualized ANC. (5)Outcomes: a composite of outcome events (feasibility, acceptability, effectiveness on perinatal outcomes, and continuum of care). (6) Effect measures: reported effect measures for the reported effect measures in the composite outcomes, either quantitatively or qualitatively synthesized, separate outcomes if available, including P-values, standard deviation, relative risk, hazard ratio, and confidence interval.

#### Outcomes and prioritization

The findings of available evidence on the feasibility of G-ANC service delivery in low-resource settings and its acceptability to pregnant women and community health workers will be synthesized. Comprehensive data on the effectiveness of G-ANC in increasing antenatal care retention and facility-based deliveries and the effect of G-ANC on perinatal outcomes and utilization of other reproductive health services will be collected. Determining the composite of perinatal outcomes and continuum of care will be considered the main outcome, while the other reported outcome events can be regarded as secondary outcomes.

#### Risk of bias assessment

The use of Joanna Briggs Institute (JBI) Critical Appraisal Tools, specifically designed for JBI Systematic Reviews of both quantitative and qualitative studies, will be used to evaluate the methodological quality of the included studies (24, 25). To assess the quality of articles for cohort, non-randomized, quasi-experimental, and randomized controlled trial (RCT) studies, the JBI Critical Appraisal Tools, which comprise specific checklists for each of the study designs, will be utilized. Two assessors (MG and HEA) will evaluate the quality of each research study, with a third reviewer (MBM) involved in resolving any discrepancies.

#### Data synthesis

The characteristics of pregnant women (number of groups, group size, number of women grouped, number of leaders in the group, who leads the group, the total number of group sessions (number of ANC), follow-up periods for the outcome and setups for G-ANC) will be assessed. Statistical heterogeneity will be assessed using the Higgins test, where the I^2^ statistics will be determined and reported. If the studies included in the analysis are homogeneous, a meta-analysis will be conducted to calculate the overall effect of G-ANC on the continuum of care and perinatal outcomes compared to conventional individualized care.

This analysis will be performed using R version 3.6.1 software, and random effects will be used to determine the weights for the meta-analysis if necessary after carefully exploring the presence and nature of heterogeneity. If there is high heterogeneity (I^2^ ≥50% or P<0.1) or if the data is incomplete, a qualitative synthesis will be conducted instead of a meta-analysis. If various types of effect measures are utilized in the original studies, such as odds ratios, risk ratios, and hazard ratios, the meta-analysis will be conducted for each type of effect measures. The study results will be reported sequentially, commencing with primary outcomes, followed by secondary outcomes and important subgroup analysis based on design, setup, country/region context, and methods of group classification. These methods will be conducted to investigate the possible causes of variability between studies and to explore the strength of the meta-analysis.

Given that the effectiveness of G-ANC can be greatly affected by the country’s context, policy, and resources, stratification will be made based on the country’s context or region (Eastern Africa, Western Africa, South Africa, and Central Africa). Additionally, study results will be stratified based on the setup where the G-ANC was conducted (facility-based vs. community-based intervention). Furthermore, study results will be stratified based on study design to assess whether the study design influences the association between exposure and outcomes. The number of women in a group and how they are grouped may impact the effectiveness of G-ANC.

For qualitative synthesis, we will put forward summary and narrative statements and quotes from the experience of pregnant women, group leaders, and health workers toward the feasibility and acceptability of G-ANC.

#### Meta-bias (es)

Outcome reporting biases will be assessed by comparing outcomes documented in research protocols with those reported in the actual study reports. Additionally, sensitivity analysis using STATA will evaluate the impact of selective reporting on the results of meta-analyses, if deemed necessary. The use of funnel plots will also be employed to investigate potential publication bias.

#### Confidence in cumulative evidence

The Grading of Recommendations, Assessment, Development, and Evaluation working group methodology will be used to assess the quality of evidence for all outcomes. The assessment will consider the domains of risk of bias, consistency, directness, precision, and publication bias. Due to the mixed nature of the studies comprising this review, including both observational and experimental research, the evidence will initially be assessed as moderate. However, if a substantial effect size is present, a dose-response relationship is established, or all potential biases are minimal, the strength of the evidence may be potentially being upgraded (26).

#### Ethics and dissemination

Ethical approval is not applicable, as no original data will be collected. The findings of this review will be disseminated through publication and conference presentations.

## Data Availability

research data will be made publicly available when the study is completed and published

## Authors Contributions

All the authors have made substantial intellectual contributions to the development of the protocol; MBM and ZHG conceptualized and designed the study. MBM and EWT formulated the search strategy. HEA developed the data synthesis section. MG and AHA drafted the risk of bias assessment section. TTC and HEA developed the meta-bias and confidence in cumulative evidence sections. TTC drafted the information sources and search strategy sections. MBM drafted the remaining sections of the protocol. ZHG and MG critically reviewed and revised the protocol. All authors read and approved the final version of the protocol. MBM serves as the guarantor of the protocol.

## Funding

This research received no specific grant from any funding agency in the public, commercial, or not-for-profit sectors.

## Competing interests

The authors declare that they have no conflicts of interest.

## Notes

### Competing Interest Statement

The authors have declared no competing interest.

### Funding Statement

The author(s) received no specific funding for this work.

### Author Declarations

Ethical approval is not applicable as no original data will be collected.

